# Multidimensional and Longitudinal Impact of a Genetic Diagnosis for Critically Ill Infants

**DOI:** 10.1101/2024.06.29.24309646

**Authors:** Monica H Wojcik, Maya C del Rosario, Henry A Feldman, Hadley Stevens Smith, Ingrid A Holm

**Author notes:** **Corresponding author:** Monica H Wojcik, Boston Children’s Hospital, 300 Longwood Ave, Boston, MA, 02115.

## Abstract

**Background and Objectives:** Many genetic conditions present in the neonatal intensive care unit (NICU), where a diagnostic evaluation is pursued. However, understanding of the impact of a genetic diagnosis on clinical outcomes and health-related quality of life for these infants remains incomplete. We therefore evaluated parent-reported outcomes complemented by clinical outcomes measures over one year for a cohort of infants in the NICU undergoing genetic evaluation.

**Methods:** Prospective cohort study evaluating outcomes after genetics consultation in a level IV NICU via parent-report and electronic medical records (EMR) review. Eligible infants were genetically undiagnosed at enrollment. Parent surveys were administered at baseline and three, six-, and 12-months following enrollment and assessed genetic testing utility as well as parent-reported infant health-related quality of life using the Infant Toddler Quality of Life Questionnaire.

**Results:** 110 infant-parent pairs were enrolled. Infants had a median age at enrollment of 15 days (interquartile range 8-37.75). At baseline, 74% (81/110) of parents endorsed high importance of finding a genetic diagnosis, but perceived importance significantly decreased over time. Over the study period, 38 infants received a molecular diagnosis per parent report, though this was discordant with EMR review. Identification of a diagnosis did not significantly impact health-related quality of life across most domains, which was lower overall than population norms.

**Conclusions:** A genetic diagnosis is highly desired by parents in the NICU, though waning interest over time for undiagnosed families may reflect parental emotional adaptation and acceptance. Additional supports are needed to improve perceived quality of life.

## Introduction

Many infants admitted to the NICU are suspected to have genetic disorders, and increased use of exome (ES) or genome sequencing (GS) has enabled precision diagnosis in a high proportion of infants tested^1-5^. However, usage and availability of diagnostic genetic testing, including ES/GS, varies widely across NICUs, even those delivering the highest levels of care^6-8^. In this setting of limited availability, decisions of whether to offer diagnostic genetic testing while in the NICU, versus deferring such testing to the outpatient setting, often take into account estimations of diagnostic and clinical utility. However, prior evaluations of clinical utility for critically ill infants have been primarily physician-centered and limited to the NICU setting, limiting the ability to understand the impact of postponing testing to after discharge^2,9,10^. This includes assessments of healthcare utilization and costs of care^11^: whereas cost-effectiveness of accelerated diagnosis via rapid ES/GS has been demonstrated from the payer or hospital perspective^3,11^, the financial impact on families remains unclear. Identifying a genetic diagnosis may serve to reduce out-of-pocket costs to families as a result of more efficient health care delivery overall, however, it is also possible that costs to families increase due to additional referrals to subspecialists after this diagnosis^11,12^.

When the parent perspective is incorporated into outcomes for NICU infants, it often centers on the utility of a testing approach, such as rapid GS, rather than the utility of a genetic diagnosis itself^13,14^. Previous studies have identified that parents are hopeful for a genetic diagnosis and identify beneficial aspects of testing, although the potential for harm to family relationships in this vulnerable time period has been raised^13-16^. Furthermore, when parent-perceived utility is assessed, it has typically been for infants enrolled in clinical trials of ES/GS, which may represent a biased population and parental surveys often do not include validated survey tools^13-16^. The impact of a genetic diagnosis on health-related quality of life for such infants is also notably absent. Thus, optimal implementation of diagnostic genetic testing in the NICU remains incompletely understood due to limited insight into of the impact of a diagnosis, particularly on longer-term health outcomes, burden of care, and infant health-related quality of life from the parent perspective. We therefore sought to explore multiple dimensions of utility in a prospective cohort of NICU infants and their parents, incorporating parent-reported outcomes complemented by clinical outcomes measures for a diverse cohort of infants in the NICU suspected to have genetic disorders over the first year of life.

## Methods

This was a prospective cohort study evaluating outcomes after genetics consultation for diagnostic evaluation in a level IV NICU (Figure 1) via parent report and electronic medical records (EMR) review.

**Figure 1.**
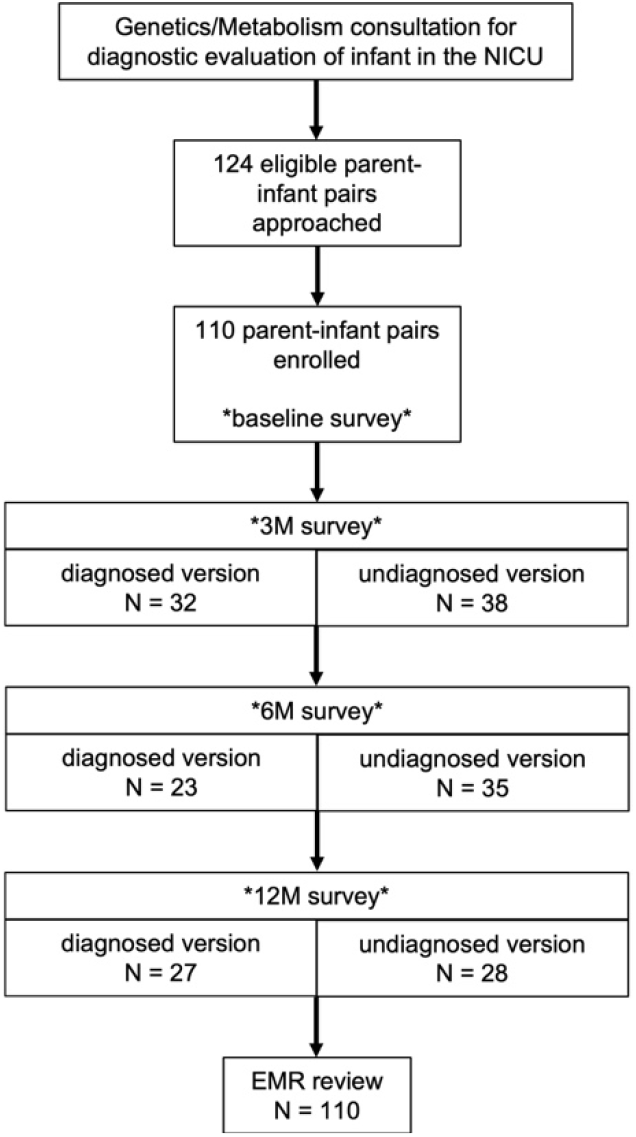
CONSORT-style depiction of study processes and retention. EMR, electronic medical records; NICU, neonatal intensive care unit; M, months. Asterisks (*) represent survey time points, with different survey versions sent to diagnosed versus undiagnosed infants (see Supplemental Figure 1).

### Study population

Infants and parents were recruited from a level IV NICU at an academic children’s hospital over a 28-month period between 2019-2021. Eligible infants were in the NICU, had undergone clinical genetics consultation, and were genetically-undiagnosed at the time of enrollment. Gift cards ($10 value) were provided to enrolled families for completion of each study survey.

### Survey design

The survey instrument was designed to explore parental values regarding a genetic diagnosis, the perceived burden of caring for their infant, and infant health-related quality of life outcomes. Cognitive interviews with medical geneticists, neonatologists, and geneticists were conducted to aid in survey development. We assessed participant demographic characteristics using items based on those used in the eMERGE network.^17^, We assessed perceived utility (Table 1) of a genetic diagnosis and burden of care using novel survey items (Appendix) that were informed by domains identified in prior qualitative work regarding the parental experience of rare disease diagnosis for a young child.^18,19^Responses were collected on a 5-point Likert scale from Not at all important (=1) to Extremely important (=5) or Strongly disagree (=1) to Strongly agree (=5). Healthcare utilization and cost to families was assessed using items adapted from surveys administered in the BabySeq Project^20^ and the National Survey of Children with Special Healthcare Needs.^21^ Health-related quality of life was evaluated using the Infant Toddler Quality of Life Questionnaire™ (ITQOL) 47-item survey, the psychometric properties of which have been validated in a population-based sample^22^ as well as in a cohort of toddlers with a history of NICU admission^23^.

**Table 1.** Demographic and clinical characteristics of study participants.

|  |  |
| --- | --- |
| **Demographics – Parent/caregiver\*** | N = 110 |
| Age, years (N = 101; median, IQR) | 32 (29-35) |
| Female, % | 84 (76%) |
| Race (multiple selections allowed) |  |
| White | 89 (81%) |
| Black | 12 (11%) |
| Asian, Hawaiian, or Pacific Islander | 5 (5%) |
| Other | 12 (11%) |
| Hispanic/Latinx (N=108) | 13 (12%) |
| Marital status |  |
| Never married | 24 (22%) |
| Married | 84 (76%) |
| Divorced | 2 (2%) |
| Living with partner (N=108) | 102 (94%) |
| Educational status |  |
| Some or all of high school | 14 (13%) |
| Some college | 25 (23%) |
| College degree | 40 (36%) |
| Post-college degree | 31 (28%) |
| Employment status (multiple selections allowed)<br>(N=109) |  |
| Currently employed | 37 (34%) |
| Temporary leave | 55 (51%) |
| Homemaker | 11 (10%) |
| Unemployed | 5 (5%) |
| Disabled | 3 (3%) |
| Student or Other | 2 (2%) |
| Income |  |
| Less than \$40,000 | 19 (17%) |
| \$40,000 to \$89,999 | 23 (21%) |
| \$90,000 to \$149,999 | 27 (25%) |
| \$150,000 or above | 27 (25%) |
| Prefer not to answer | 14 (13%) |
| Insurance (N = 108) |  |
| Employer | 80 (74%) |
| MassHealth | 25 (23%) |
| Both | 3 (3%) |
| **Demographics – Infant** | N = 110 |
| Female, % | 50 (46%) |
| Preterm, % | 43 (39%) |
| Gestational age on admission (if preterm) |  |
| 24 weeks or less | 2 (5%) |
| 28-31 completed weeks | 5 (12%) |
| 32-33 completed weeks | 18 (42%) |
| 34-37 | 18 (42%) |
| Age at NICU admission, days (median, IQR) | 7.5 (2-29.75) |
| Age at enrollment, days (median, IQR) | 15 (8 – 37.75) |
| Age at genetics consult (median, IQR) | 9 (4 – 26.75) |
| Initial consult team |  |
| Genetics | 76 (69%) |
| Metabolism | 34 (31%) |
| Critical illness at enrollment** | 56 (51%) |

IQR, interquartile range; NICU, neonatal intensive care unit; \*all but one survey respondent identified themselves as a biological parent; \*\*defined as need for respiratory or hemodynamic support

Following the baseline survey, follow-up surveys were identical for all participants with regards to questions involving clinical outcomes, burden of care, and health-related quality of life. They differed only in the phrasing of questions related to parent-perceived utility of a genetic diagnosis, where undiagnosed families continued to receive the same questions presented at baseline regarding the hypothetical value of a diagnosis (Table 1), while diagnosed families (diagnosed status ascertained per parent report) received similar questions regarding the actual value of the diagnosis received.

### Data collection

The baseline study survey was completed by a parent in the NICU immediately following enrollment. and follow-up surveys were sent at 3-, 6-, and 12-months after enrollment. For all enrolled infants, data pertaining to clinical features and diagnoses, diagnostic outcomes, and healthcare utilization were abstracted from the EMR and reviewed by two independent investigators (MHW, MCD) for accuracy.

### Analysis

Statistical analyses were conducted in R^24^, including descriptive statistics and mixed-effects linear regression models on the primary outcomes of 1) standardized ITQOL scores and 2) parental responses to items scored on a 5-point Likert scale. ITQOL raw scores were translated into standardized scores (minimum 0 to maximum 100) as per the ITQOL manual^25^ and compared to population normative values provided by the survey developer^26^. The Wilcoxon test was used for comparisons of continuous outcomes and a Fisher’s exact test to compare proportions, with *p* < 0.05 considered statistically significant.

## Results

Of 124 eligible infant-parent pairs approached, and 110 enrolled (89%) and all 110 completed the baseline survey (Figure 1). Demographics of infants and caregivers at baseline are presented in Table 1. Demographic factors did not vary substantially between parents who completed or did not complete follow-up surveys, with the exception of gender and age; male caregivers were less likely to complete surveys at the 3-month timepoint compared to the baseline survey (OR = 0.24 95% CI 0.06-0.82 *p* = 0.027) and odds of survey completion were lower with increasing parental age (OR = 0.88 95%CI 0.77-0.99, *p* = 0.046).

### Diagnostic Utility

#### Electronic medical record

The primary reason for genetics consultation included: multiple congenital anomalies (40/110, 36%), single anomaly (21/110, 19%), complex metabolic (14/110, 13%), hypotonia (10/110, 9%), abnormal newborn screen (10/110, 9%), seizure (5/110, 5%), and other infrequent indications (10/110, 9%). Over the study period, 42/110 (38%) infants had molecular diagnoses identified per EMR review, with 38/78 (49%) parents who completed at least one follow-up survey reporting their infant as diagnosed within this same timeframe. Per EMR review, median age of diagnosis was 36 days (IQR 22-66.5 days), and 25/42 (59%) of molecular diagnoses were returned while the infant was in the hospital. Test leading to diagnosis included chromosomal microarray (5/42, 12%), single gene sequencing (6/42, 14%), gene panel testing (10/42, 24%), exome sequencing (19/42, 45%) and other tests in one infant each: somatic cytogenetics and Beckwith-Wiedemann methylation analysis.

#### Parent report

Concordance between diagnosed status per EMR review versus parental perception was high but not complete. Two parents with molecular diagnoses assessed by EMR review reported their infant undiagnosed: one who was found to have carrier status for a genetic metabolic condition explaining the abnormalities that prompted the genetic testing and one with a variant of uncertain significance (VUS) identified via exome sequencing that was felt by specialist providers to explain the infant’s rare immunologic presentation. Another 9 parents with diagnosed infants did not complete follow-up surveys, in five cases because the infant died soon after the initial survey, thus 2/32 (6%) diagnosed infants whose parents completed follow up surveys were reported as undiagnosed. Conversely, eight parents reported that their infant was diagnosed with a genetic disorder on the survey when EMR review did not reveal a genetic diagnosis: three parents had infants with recurring constellations of embryonic malformations (heterotaxy, VACTERL association, hemifacial microsomia, Femoral Facial Syndrome), one had hypotonia and feeding difficulty of unknown cause, one had cardiac dysfunction and concern for genetic metabolic disorder, and the remaining two parents reversed their impression of diagnosed status on subsequent follow-ups surveys.

### Clinical Utility

#### Electronic Medical Record

Health outcomes ascertained by EMR review are presented in **Table 2**. Across the cohort, 14/110 (13%) infants died within the first year of life. Over the first year of life, 108/110 infants in the cohort (98%) had at least one subspecialty consultation in addition to the genetics or metabolism consultation; the median number of consultations was five, with a range from zero to fifteen (IQR 3-8) (the two infants with no specialist consultations were evaluated for abnormal newborn screens by the metabolism team). The median number of outpatient subspecialty clinic visits in our EMR was 7 (IQR 3-9), with the caveat that not all infants continued to receive their outpatient care within our institution, or may have received care at more than one institution. Most infants (73/110, 66%) had at least one surgery in the first year of life, and some infants had multiple admissions to our NICU, with a median number of admissions per infant of 1 (IQR 1-2, maximum 4). The total length of stay of the first admission to BCH, including time spent in the NICU as well as elsewhere, was median of 27 days (IQR 12-42 days) and total number of days spent in our NICU was a median of 17 (IQR 10-36). In the first year of life, the total number of hospital days at our institution across the cohort was a median of 36 (IQR 20-67.5). There were no differences in these outcomes comparing the diagnosed to undiagnosed infants (defined by molecular diagnosis identified upon EMR review, **Table 2**).

**Table 2.**
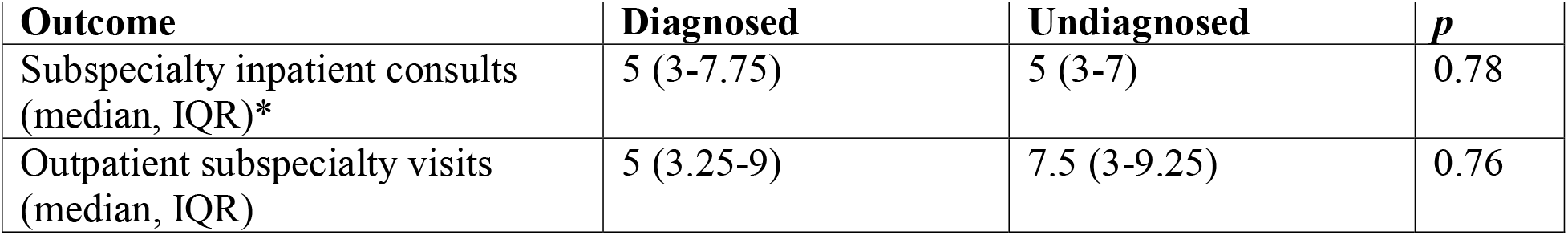
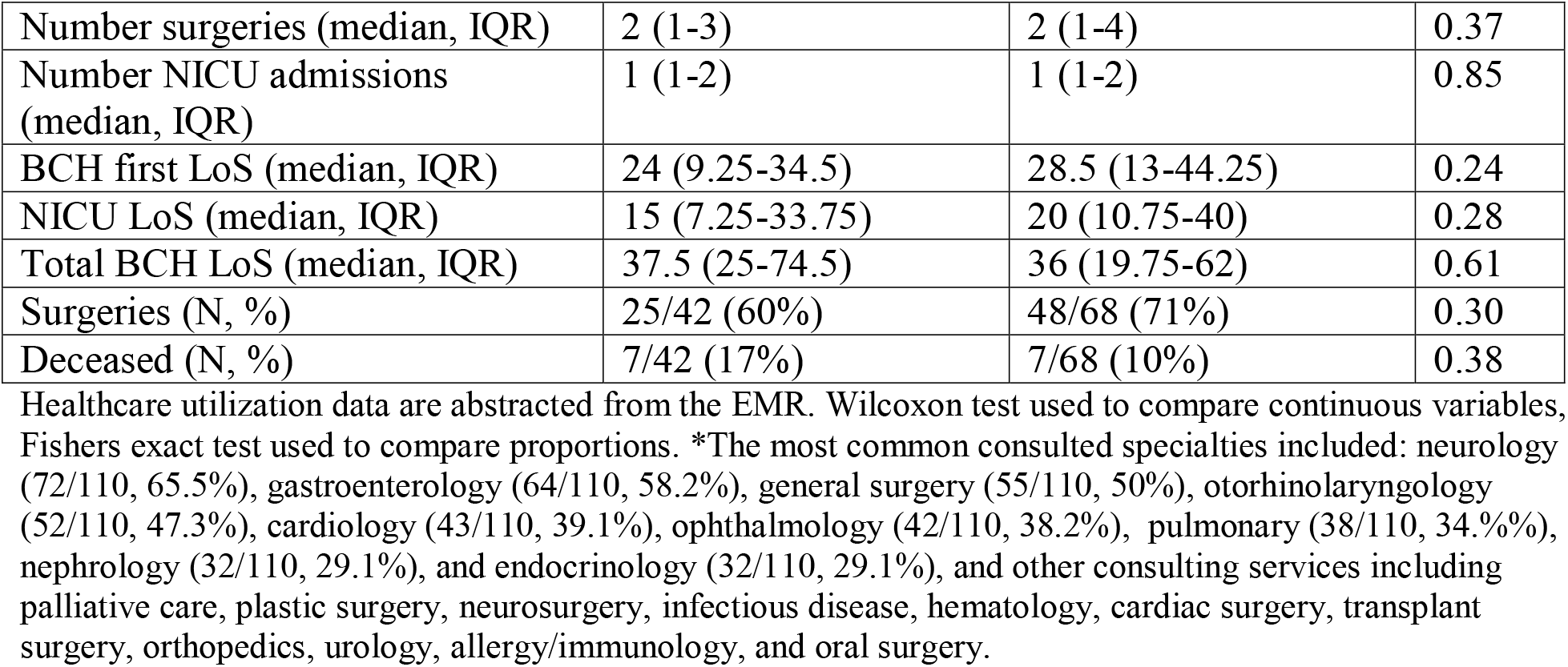
Impact of a diagnosis on healthcare utilization.

| **Table 2. Impact of a diagnosis on healthcare utilization** |  |  |  |
| --- | --- | --- | --- |
| **Outcome** | **Diagnosed** | **Undiagnosed** | ***p*** |
| Subspecialty inpatient consults (median, IQR)* | 5 (3-7.75) | 5 (3-7) | 0.78 |
| Outpatient subspecialty visits (median, IQR) | 5 (3.25-9) | 7.5 (3-9.25) | 0.76 |
| Number surgeries (median, IQR) | 2 (1-3) | 2 (1-4) | 0.37 |
| Number NICU admissions (median, IQR) | 1 (1-2) | 1 (1-2) | 0.85 |
| BCH first LoS (median, IQR) | 24 (9.25-34.5) | 28.5 (13-44.25) | 0.24 |
| NICU LoS (median, IQR) | 15 (7.25-33.75) | 20 (10.75-40) | 0.28 |
| Total BCH LoS (median, IQR) | 37.5 (25-74.5) | 36 (19.75-62) | 0.61 |
| Surgeries (N, %) | 25/42 (60%) | 48/68 (71%) | 0.30 |
| Deceased (N, %) | 7/42 (17%) | 7/68 (10%) | 0.38 |

Healthcare utilization data are abstracted from the EMR. Wilcoxon test used to compare continuous variables, Fishers exact test used to compare proportions. \*The most common consulted specialties included: neurology (72/110, 65.5%), gastroenterology (64/110, 58.2%), general surgery (55/110, 50%), otorhinolaryngology (52/110, 47.3%), cardiology (43/110, 39.1%), ophthalmology (42/110, 38.2%), pulmonary (38/110, 34.4%), nephrology (32/110, 29.1%), and endocrinology (32/110, 29.1%), and other consulting services including palliative care, plastic surgery, neurosurgery, infectious disease, hematology, cardiac surgery, transplant surgery, orthopedics, urology, allergy/immunology, and oral surgery.

#### Parent report

Parent-reported health outcomes across the 12-month follow-up period were also similar between diagnosed and undiagnosed infants, as was use of Early Intervention services, number of outpatient clinic visits, number of surgical procedures, number of emergency department or urgent care visits, and number of nights in the hospital or intensive care unit. The one exception was at the three-month timepoint, where a lower proportion of undiagnosed infants had no services (6/38, 16%) compared to diagnosed infants (15/32, 47%). Of note, even at baseline, 41/110 infants had already had at least one surgery, 23/110 had been home since birth, and 22/110 had been to the emergency department (**Supplemental Table 1**).

In terms of parent-reported financial and economic impact, at baseline, half of parents had stopped work or cut down on their hours to care for their infant, and 25.5% reported financial problems as a result of their infant’s care. At 12 months, 30% of diagnosed and 32% of undiagnosed families reported financial problems, with no significant difference by diagnosed status. Many parents (close to half at all time points) reported that they had cut down their hours at work due to the infant’s medical issues, although there was no significant difference by diagnosed status. Most parents reported at least some out-of-pocket costs for medical care, with 40% of diagnosed and 29% of undiagnosed parents reporting that they had spent more than $5000 in out-of-pocket costs at the 12-month time point (*p* = 0.52). (**Supplemental Table 2**)

### Personal Utility

At baseline, 74% (81/110) of parents felt that identifying a genetic diagnosis was very or extremely important. The perceived importance of a diagnosis for families that remained undiagnosed decreased over the survey timepoints (**Figure 2**), and a mixed effects linear regression model evaluating the impact of time (in months) on perceived importance on a 5-point Likert scale revealed a significant negative association (ß = -0.09± 0.018, *p* < 0.001, **Table 3)**.

**Table 3.**
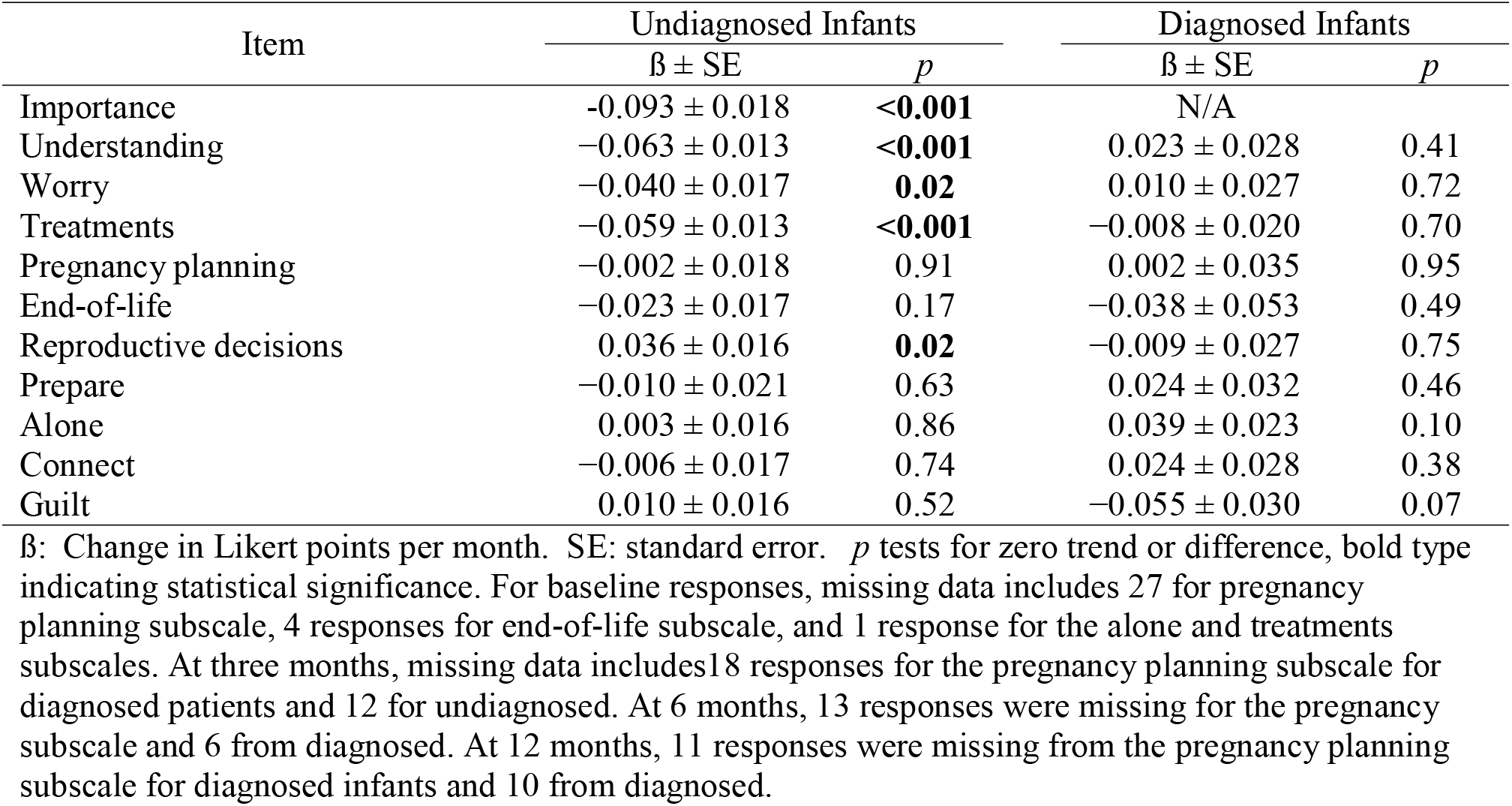
Impact of time on perceived utility of a genetic diagnosis for undiagnosed and diagnosed infants.

| Item | Undiagnosed Infants |  | Diagnosed Infants |  |
| --- | --- | --- | --- | --- |
| | $\beta \pm SE$ | *p* | $\beta \pm SE$ | *p* |
| Importance | -0.093 ± 0.018 | **<0.001** | N/A | |
| Understanding | -0.063 ± 0.013 | **<0.001** | 0.023 ± 0.028 | 0.41 |
| Worry | -0.040 ± 0.017 | **0.02** | 0.010 ± 0.027 | 0.72 |
| Treatments | -0.059 ± 0.013 | **<0.001** | -0.008 ± 0.020 | 0.70 |
| Pregnancy planning | -0.002 ± 0.018 | 0.91 | 0.002 ± 0.035 | 0.95 |
| End-of-life | -0.023 ± 0.017 | 0.17 | -0.038 ± 0.053 | 0.49 |
| Reproductive decisions | 0.036 ± 0.016 | **0.02** | -0.009 ± 0.027 | 0.75 |
| Prepare | -0.010 ± 0.021 | 0.63 | 0.024 ± 0.032 | 0.46 |
| Alone | 0.003 ± 0.016 | 0.86 | 0.039 ± 0.023 | 0.10 |
| Connect | -0.006 ± 0.017 | 0.74 | 0.024 ± 0.028 | 0.38 |
| Guilt | 0.010 ± 0.016 | 0.52 | -0.055 ± 0.030 | 0.07 |

**Figure 2.**
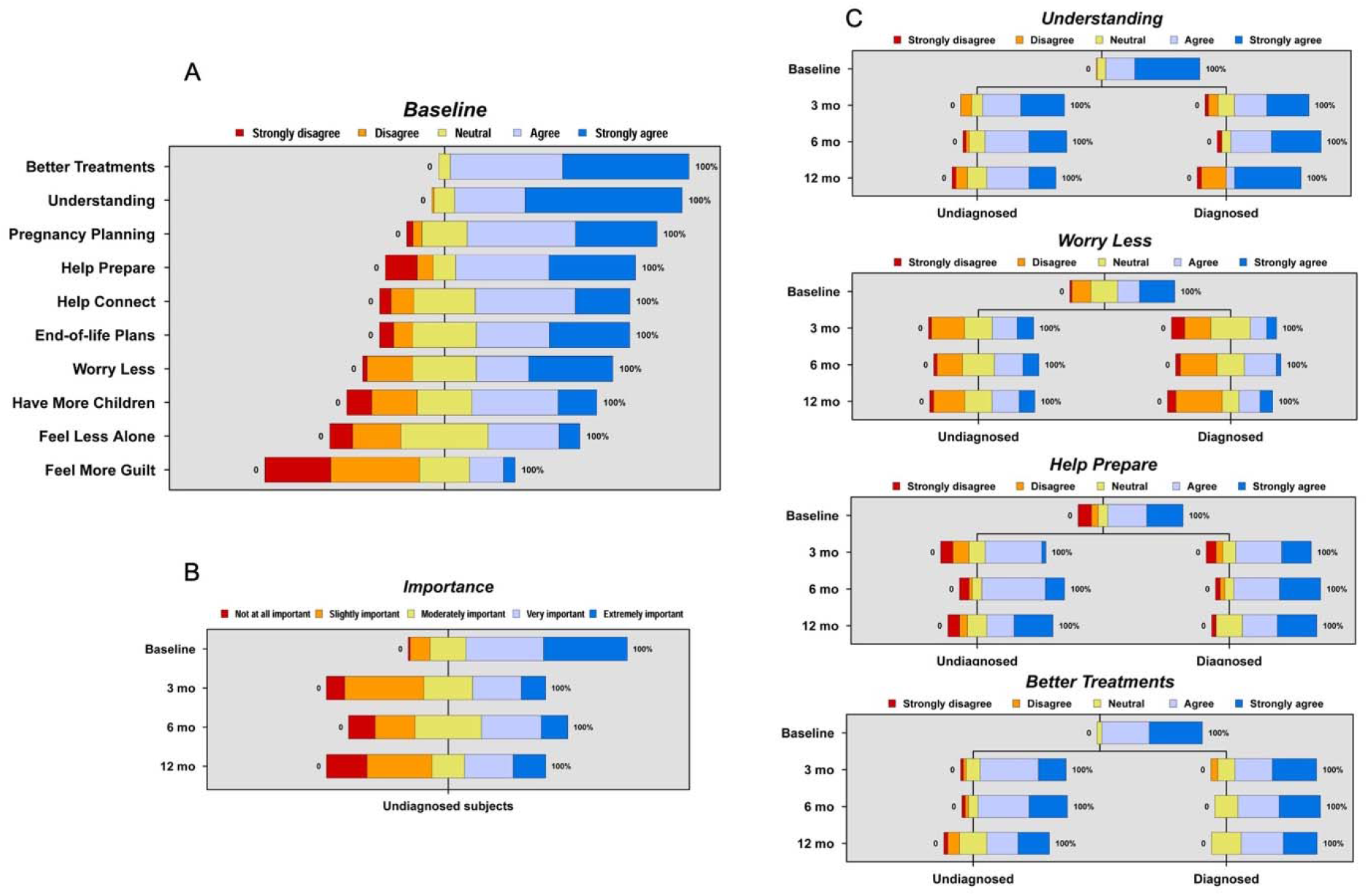
Parent-reported values at baseline and over time. Plots depict proportions of responses to a 5-point Likert scale related to importance of a diagnosis at Baseline (A), change in perceived importance for undiagnosed infants over time (B), and changes over time for diagnosed and undiagnosed infants for significant constructs (C).

At baseline, parents also endorsed the most agreement with statements that a genetic diagnosis would aid in finding better treatments for their infants (agree or strongly agree, N = 104/110, 95%) followed by improved understanding (agree or strongly agree, N = 100/110, 91%). Time (estimated by timepoint of survey completion) had a significant and negative impact on level of agreement that a diagnosis would aid in finding better treatments, in understanding and helping parents to worry less, and a significantly positive association with the perceived impact of a diagnosis on reproductive decision-making (**Figure 2, Table 3**).

Differences in the impact of time on these costructs were also identified between diagnosed and undiagnosed families. The majority of parents whose infants were diagnosed at the 3-month timepoint agreed that the diagnosis aided in understanding their infant’s medical problems (23/32, 72%), improved feelings of preparation (23/32, 72%), and helped identify better treatments for their infant (24/32, 75%), whereas a minority reported that the diagnosis helped them to worry less (8/32, 25%), or feel less alone (12/32, 38%) (**Supplemental Table 3**). These feelings remained stable, with no significant impact of time noted for any domain (**Table 4)**.

**Table 4.** Impact of time and diagnosed status on infant quality of life.

| Subscale | Time |  | Diagnosis |  |
| --- | --- | --- | --- | --- |
| | $\beta \pm SE$ | *p* | $\beta \pm SE$ | *p* |
| Overall health | 0.72 ± 0.32 | **0.03** | -7.94 ± 4.85 | 0.10 |
| Growth and development | -0.25 ± 0.33 | 0.44 | -3.46 ± 3.87 | 0.37 |
| Discomfort and pain | -0.00 ± 0.31 | 0.99 | 6.78 ± 4.09 | 0.10 |
| Temperament and mood | 0.15 ± 0.26 | 0.55 | 1.82 ± 2.86 | 0.53 |
| General health perception | -0.17 ± 0.22 | 0.45 | -1.23 ± 3.45 | 0.72 |
| Parental impact: emotional | 0.43 ± 0.37 | 0.25 | -3.37 ± 5.04 | 0.51 |
| Parental impact: time | 0.62 ± 0.31 | **0.05** | -1.36 ± 4.02 | 0.76 |
| Family cohesion | 0.17 ± 0.32 | 0.60 | 0.52 ± 4.36 | 0.91 |

β: Change in ITQOL standardized score per month, adjusted for diagnosed status; or difference in ITQOL standardized score between diagnosed and undiagnosed, adjusted for time. SE: standard error. *p* tests for zero trend or difference, bold type indicating statistical significance.

When taking into consideration the type of consultation the infants had in the NICU (genetics vs metabolism), perceived importance of a genetic diagnosis at three months was higher for infants with metabolism consults compared to those with genetics consultations (*p* = 0.04) but not at the 12-month timepoint (p = 0.20). Parents of infants with metabolism consultations also had significantly higher perceived importance of a confirmed genetic diagnosis at 12-month on finding better treatments (*p* = 0.001) and preparing for the future (*p* = 0.002) (**Supplemental Table 4**).

Quality of life, reflected in mean standardized scores (range: 0-100) from the ITQOL, did not differ between diagnosed and undiagnosed infants across all subdomains with the exception of discomfort and pain at 3 months (63.5 vs 74.2, *p* = 0.04), and general health perception at 12 months (55.3 vs 43.0 *p* = 0.008) for undiagnosed versus diagnosed infants, respectively (**Supplemental Table 5**). Across the entire cohort, quality of life was lower at 12 months for all quality of life subscales compared to population norms for healthy infants with the exception of the “temperament and mood” and “family cohesion” subscales, and was lower for all subscales except for “combined behavior” and “family cohesion” compared to norms for infants with multiple chronic conditions. In a series of multivariate mixed-effects linear regression modes incorporating both time and parent-reported diagnosed status (which may have changed over time), time had a variable impact across domains for both diagnosed and undiagnosed infants. Both the overall health and parental time impact subscales significantly increased over time, although diagnosis continued to have no significant impact on this outcome (**Table 4**).

Finally, a multivariable linear regression model (**Table 5)** was created for both the 3-month and 12-month timepoints to evaluate the impact of diagnosed status (per parental report) and potential confounders of prematurity and genetic vs metabolic disease on health-related quality of life outcomes. Overall health and general health perception were significantly lower for diagnosed infants, accounting for prematurity and type of consultation.

**Table 5.** Impact of multiple factors on infant quality of life at 3- and 12-month timepoints.

| Subscale | Diagnosis |  | Consult Type |  | Prematurity |  |
| --- | --- | --- | --- | --- | --- | --- |
|  | β ± SE | *p* | β ± SE | *p* | β ± SE | *p* |
| *3-month survey* |  |  |  |  |  |  |
| Overall health | 5.11 ± 6.34 | 0.43 | 12.73 ± 7.09 | 0.08 | 0.70 ± 6.86 | 0.92 |
| Growth/development | 0.24 ± 4.42 | 0.96 | 6.54 ± 4.93 | 0.19 | 7.47 ± 4.77 | 0.12 |
| Discomfort and pain | 11.10 ± 5.14 | **0.03** | 7.57 ± 5.74 | 0.19 | -4.48 ± 5.56 | 0.42 |
| Temperament/mood | 0.04 ± 4.17 | 0.99 | 3.18 ± 4.67 | 0.50 | 3.24 ± 4.51 | 0.48 |
| General health perception | 0.15 ± 5.09 | 0.98 | -0.47 ± 5.69 | 0.93 | 3.95 ± 5.50 | 0.48 |
| Parental impact: emotional | -3.02 ± 6.62 | 0.65 | 7.57 ± 7.40 | 0.31 | 8.15 ± 7.16 | 0.26 |
| Parental impact: time | 5.02 ± 5.45 | 0.36 | 7.25 ± 6.09 | 0.24 | 1.39 ± 5.89 | 0.81 |
| Family cohesion | 1.00 ± 6.14 | 0.87 | 2.02 ± 6.86 | 0.77 | -0.58 ± 6.64 | 0.93 |
| *12-month survey* |  |  |  |  |  |  |
| Overall health | -19.60 ± 6.80 | **0.006** | -1.75 ± 7.92 | 0.83 | -2.01 ± 7.77 | 0.80 |
| Growth/development | -9.77 ± 6.24 | 0.124 | -0.79 ± 7.27 | 0.92 | 6.53 ± 7.12 | 0.36 |
| Discomfort and pain | 6.12 ± 6.33 | 0.34 | -2.62 ± 7.38 | 0.72 | -7.54 ± 7.23 | 0.30 |
| Temperament/mood | 4.78 ± 4.27 | 0.27 | -10.07 ± 4.97 | **0.05** | 3.83 ± 4.87 | 0.44 |
| General health perception | -12.04 ± 4.62 | **0.01** | -2.92 ± 5.38 | 0.59 | -0.96 ± 5.28 | 0.86 |
| Parental impact: emotional | -7.16 ± 7.20 | 0.33 | -2.34 ± 8.39 | 0.78 | 5.65 ± 8.23 | 0.50 |
| Parental impact: time | -3.67 ± 6.40 | 0.57 | 1.04 ± 7.46 | 0.89 | 2.40 ± 7.32 | 0.75 |
| Family cohesion | 5.87 ± 5.43 | 0.29 | -11.67 ± 6.33 | 0.07 | 11.47 ± 6.20 | 0.07 |

β: Difference in ITQOL standardized score attributed to diagnosis, metabolism consult service versus genetics consult service (reference), or prematurity. SE: standard error. *p* tests for zero difference, bold type indicating statistical significance.

## Discussion

For a cohort of NICU infants suspected to have genetic disorders, we found that a genetic diagnosis is highly desired by parents and motivated by potential for improved understanding, preparation, and treatments – particularly for infants with suspected inborn errors of metabolism. However, the perceived importance of a genetic diagnosis and its perceived utility waned over time for families who remained undiagnosed. In addition, parent-reported infant health-related quality of life was lower across nearly all domains for our cohort compared to population norms. We found no lack of significant impact of a genetic diagnosis on health-related quality of life over the study period for these infants.

Our findings highlight the complexities in assessing the clinical and perceived utility of a genetic diagnosis for infants admitted to the NICU, where prior studies have identified high clinical utility from the physician perspective^1,2^ and prior evaluations of parent-reported outcomes have reported high perceived value across multiple domains^14-16^. However, these prior studies have focused on infants are enrolled in genomic sequencing studies (ES or GS)^14-16^ which may introduce selection bias, as parents who enroll in a prospective sequencing study may have different views than those receiving usual care – particularly as they have presumably had the potential benefits of a diagnosis explained to them as part of the study enrollment process. This may explain why we did not find as clear of an endorsement across domains such as preparation for the future and forming connections^16^ for infants who were diagnosed over our study period and why we did not find negative impact on family functioning as has been previously suggested^15^. Our finding of discordance of diagnosed status between parent report compared to the EMR is also notable, replicates findings in prior studies^14,27^, and stresses the importance not only of assessing parent-reported outcomes but also of improved post-results genetic counseling.

It is also important to note that perceived utility in our study varied by presenting phenotype, reflected in our comparison of infants who had consultations for metabolic conditions to those with consultations for other genetic syndromes. This may be attributed to more concrete clinical management changes with tangible benefits seen for metabolic conditions: for example, inborn errors of metabolism that may present in the NICU and are often managed by dietary or medication changes that directly impact illness trajectory. Indeed, the parents in our cohort whose infants had consultations for metabolic disorders perceived a higher utility of their diagnoses regarding the impact on treatments compared to those with general genetics consultations. However, having a genetics versus a metabolism consult did not impact perceived quality of life, suggesting that treatment impact alone may not influence this outcome.

Decreased perceived importance and value in a diagnosis over time for undiagnosed infants may reflect parental emotional adaptation and acceptance. Indeed, it is notable that of all the subscales in the ITQOL instrument, “family cohesion” was consistently not lower in our cohort compared to population norms, as other subscales were, and also did not significantly differ between diagnosed and undiagnosed infants. This is in contrast to a prior study surveying parents 12 weeks or more after rapid ES in the NICU, where family functioning was lower in diagnosed families compared to undiagnosed, raising the concern that rapid genomic sequencing may be harmful for families^15^. Our results instead suggest preservation of family functioning that may contribute to emotional adaptation. Conversely, while perceived importance for undiagnosed infants decreased over time, agreement across all utility items remained constant for infants who were diagnosed, supporting the durability of benefits of a genetic diagnosis. The lack of impact of a genetic diagnosis on quality of life at the 12-month timepoint may reflect the competing effects of severity of illness in these infants against the psychosocial benefits of diagnostic clarity; indeed, prior work has suggested that parental expectations of genetic diagnostic testing are not always matched by reality^28^.

Limitations to our study relate to sample size, although our sample size is similar to prior analyses of this patient population^14,15^. Larger cohorts are ultimately needed to capture a wide variety of phenotypes with differential impacts. Nonetheless, these data present an important glimpse into uncharted territory regarding parent-perceived utility and healthcare utilization in a cohort unbiased by selection for enrollment in a clinical sequencing study. Furthermore, defining these dimensions of utility over the natural course of usual care benchmarks outcomes for future NICU sequencing studies.

Overall, our results illustrate a high perceived importance of a genetic diagnosis to families of critically ill infants. That this perceived importance wanes over time supports the need for broad diagnostic testing in the moment that it is thought to be most useful for families: while in the NICU. Attempts to defer testing to the outpatient setting, by which time the infant may be 6-12 months old, are more vulnerable to missed care opportunities as parental interest wanes and the focus may be shifting to day-to-day management of infants with medical complexity. Our study further suggests that there is a need for improved family-centered longitudinal support to address health-related quality of life and supports the importance of parent-reported quality of life measures, particularly as parental perceptions may substantially differ from physician-perceived quality of life^29^. While genetic diagnosis, often achieved in the NICU via rapid genomic sequencing, holds value, the exact nature of this value remains unclear. While we and others have shown that healthcare utilization may not be substantially impacted by a genetic diagnosis^11,30^, parent-perception of care burden may be improved. Given the dynamic nature of perceived utility, implications for health-related quality of life, and impact of infant phenotype, increased incorporation of longitudinal and multidimensional parent-reported outcomes in genomic sequencing trials is critical to best define its optimal implementation.

## Data Availability

All data produced in the present study are available upon reasonable request to the authors

## Abbreviations

EMR: electronic medical record
ITQOL: infant-toddler quality of life questionnaire
IQR: interquartile range
NICU: neonatal intensive care unit

