## Supplemental Table 1 for "Multidimensional and Longitudinal Impact of a Genetic Diagnosis for Critically Ill Infants"

| **Supplemental Table 1. Healthcare utilization per parent report** | | | | | | | |
| --- | --- | --- | --- | --- | --- | --- | --- |
|  | **Baseline** | **3 months** | | **6 months** | | **12 months** | |
|  |  | Diagnosed  N = 32 | Undiagnosed  N = 38 | Diagnosed  N = 23 | Undiagnosed  N = 35 | Diagnosed | Undiagnosed |
| Home from hospital? | 23/110 (20.9%) | 26/32 (81%) | 33/38 (87%)  *p* = 0.74 | 21/22 (96%) | 32/33 (97%)  *p* = 1.0 | 25/25 (100%) | 27/27 (100%) |
| Early Intervention*  Yes  No & not planning  No & planning | 1/22 (5%)  10/22 (45%)  11/22 (50%) | 14/25 (56%)  4/25 (16%)  7/25 (28%) | 26/33 (79%)  5/33 (15%)  2 (6%)  *p* = 0.08 | 17/21 (81%)  2/21 (10%)  2/21 (10%) | 25/32 (78%)  3/32 (9%)  4/32 (13%)  *p* = 1.0 | 22/25 (88%)  3/25 (12%)  0 (0%) | 24/27 (89%)  2/27 (7%)  1/27 (4%)  *p* = 0.83 |
| EI hours  <1  1-3  4-6  6-10  Over 10 |  | 5/14 (36%)  8/14 (57%)  0  1/14 (7.1%)  0 | 11/26 (42%)  14/26 (54%)  1/26 (4%)  0/26 (0%)  0  *p* = 0.47 | 7/17 (41%)  7/17 (41%)  3/17 (18%)  0  0 | 10/25 (40%)  14/25 (56%)  1/25 (1%)  0  0  *p* = 0.38 | 8/22 (36%)  13/22 (59%)  0  0  1/22 (5%) | 5/24 (21%)  18/24 (75%)  0  0  1/24 (4%)  *p* = 0.66 |
| EI service types  PT  OT  Speech  Developmental  None |  | 7/32 (22%)  5/32 (16%)  5/32 (16%)  5/32 (16%)  15/32 (47%) | 20/38 (53%)  11/38 (29%)  23/38 (32%)  5/38 (13%)  6/38 (16%)** | 8/23 (35%)  6/23 (26%)  9/23 (39%)  6/23 (26%)  9/23 (39%) | 20/35 (57%)  10/35 (29%)  8/35 (23%)  4/35 (11%)  7/35 (20%) | 17/27 (63%)  11/27 (41%)  11/27 (41%)  9/27 (33%)  6/27 (22%) | 19/27 (70%)  11/27 (41%)  12/27 (44%)  6/27 (22%)  5/27 (19%) |
| EI service plans  PT  OT  Speech  Developmental  None/all set | 4/23  5/23  4/23  3/23  10/23 | 8/32 (25%)  8/32 (25%)  9/32 (28%)  12/32 (38%)  12/32 (38%) | 9/38 (24%)  7/38 (18%)  8/38 (21%)  9/38 (24%)  16/38 (42%) | 5/23 (22%)  5/23 (22%)  4/23 (17%)  5/23 (22%)  16/23 (70%) | 5/35 (14%)  6/35 (17%)  9/35 (26%)  7/35 (20%)  17/35 (49%) | 7/27 (26%)  6/27 (22%)  9/27 (33%)  5/27 (19%)  11/27 (41%) | 3/27 (11%)  5/27 (19%)  8/27 (30%)  6/27 (22%)  11/27 (41%) |
| Outpatient visits  (median, IQR) | 1 (0-3) | 4 (3-10)  N = 12 | 7 (3.75-10)  N = 18  *p* = 0.21 | 10 (5.25-15)  N = 18 | 10 (6-20)  N = 24  *p* = 0.81 | 15 (7-25)  N = 21 | 17.5 (8.5-28.75)  N = 22  *p* = 0.77 |
| Specialist visits  (median, IQR) | 3.5 (1-5.25)  N = 8 | 5 (4.25-7.5)  N = 21 | 3 (2-4.75)  N = 27  *p* = 0.13 | 8 (7-20)  N = 17 | 7.5 (5-20)  N = 22  *p* = 0.38 | 10 (10-20)  N = 21 | 10 (7.25-20)  N = 22  *p* = 0.91 |
| ER/Urgent care visits*  Yes | 21/22 (95%) | 18/26 (69%) | 17/32 (53%)  *p* = 0.28 | 14/21 (67%) | 18/32 (56%)  *p* = 0.57 | 7/24 (29%) | 13/27 (48%)  *p* = 0.25 |
| Number of ER/Urgent*** | n/a | 1 (1-2.75)  N = 18 | 1 (1-3)  N = 17  *p* = 0.26 | 3 (1-5)  N = 13 | 3 (1.25-3)  N = 18  *p* = 0.60 | 4 (2-6)  N = 17 | 4 (3-5)  N = 14  *p* = 0.98 |
| Hospital nights | 14 (7-31)  [N=108] | 35 (17-94.5)  N = 31 | 51 (29-90)  N = 37  *p* = 0.51 | 74.5 (26-141)  N = 20 | 50 (28.5-68.5)  N = 31  *p* = 0.22 | 40 (27.5-82)  N = 23 | 39.5 (22-60)  N = 22  *p* = 0.72 |
| ICU nights | 13 (6-28.5)  [N=106] | 30 (14-79.5)  N = 31 | 43 (20.25-63.75)  N = 38  *p* = 0.36 | 26 (14.25-87)  N = 22 | 42 (15.5-60)  N = 31  *p* = 0.91 | 19 (10.75-48.75)  N = 24 | 28 (10-59)  N = 24  *p* = 0.67 |
| Has had surgery | 41/109 (37.6%) | 18/32 (56%) | 24/38 (63%)  *p* = 0.62 | 16/22 (73%) | 23/33 (70%)  *p* = 1.0 | 14/24 (58%) | 21/27 (78%)  *p* = 0.23 |
| Number of surgeries | 1 (1-2, range 1-4) [n=109] | 2 (2-4)  N = 18 | 2 (1-3.25)  N = 24  *p* = 0.46 | 2 (1.75-3.25)  N = 16 | 2 (1.5-3)  N = 23  *p* = 0.99 | 2 (1.25-3)  N = 14 | 3 (2-4)  N = 19  *p* = 0.28 |

*only if home; ***p* = 0.008; ***free text, if reported “30+” used 30, if reported 10-20 used lower number.

| **Supplemental Table 2** | | | | | | | |
| --- | --- | --- | --- | --- | --- | --- | --- |
|  | **Baseline** | **3 months** | | **6 months** | | **12 months** | |
|  |  | Diagnosed | Undiagnosed | Diagnosed | Undiagnosed | Diagnosed | Undiagnosed |
| Medical care costs  More than 5000  Less than 5000  Nothing  Don’t know | 9/110 (8.1%)  13/110 (11.8%)  67/110 (61%) | 11/23 (48%)  10/23 (44%)  2/23 (9%)  9 | 8/35 (23%)  20/35(57%)  7/35 (20%)  2  *p* = 0.14 | 6/19 (32%)  11/19 (58%)  2/19 (11%)  3 | 10/32 (31%)  19/32 (59%)  3/32 (9%)  1  *p* = 1.0 | 8/20 (40%)  12/20 (60%)  0/20 (0%)  5 | 7/24 (29%)  15/24 (63%)  2/24 (8%)  3  *p* = 0.52 |
| Financial problems  Yes  No  I don’t know | 28/110 (25.5%)  40/110 (36.4%)  42/110 (38.2%) | 11/32 (34%)  12/32(38%)  9/32 (28%) | 14 (37%)  20 (53%)  4 (11%)  *p* = 0.14 | 7/22 (32%)  10/22 (46%)  5/22 (23%) | 17/33 (52%)  12/33 (36%)  4/33 (12%)  *p* = 0.31 | 8/27 (30%)  17/27 (63%)  2/27 (7%) | 8/25 (32%)  13/25 (52%)  4/25 (16%)  *p* = 0.52 |
| Stop work  Yes  No  I don’t know | 55/110 (50.0%)  48/110 (43.6%)  7/110 (6.4%) | 14/30 (47%)  16/30 (53%)  2 | 16/38 (42%)  22/38 (58%)  *p* = 0.81 | 9/21 (43%)  12/21 (57%)  1 | 8/32 (25%)  24/32 (75%)  1  *p* = 0.23 | 9/25 (36%)  16/25 (64%) | 6/26 (23%)  20/26 (77%)  1  *p* = 0.36 |
| Cut down hours  Yes  No  I don’t know | 56/110 (50.9%)  47/110 (42.7%)  7/110 (6.4%) | 13/29 (45%)  16/29 (55%)  3 | 24/38 (63%)  14/38 (37%)  *p* = 0.15 | 9/21 (43%)  12/21 (57%)  1 | 17/32 (53%)  15/32 (47%)  1  *p* = 0.58 | 15/25 (60%)  10/25 (40%) | 12/26 (46%)  14/26 (54%)  1  *p* = 0.40 |
| Care coordination  More than 1 hour  Less than 1 hour  Around the clock  Don’t know | n/a | 15/32 (47%)  7/32 (22%)  4/32 (13%)  6/32 (19%) | 12/37 (32.4%)  8/37 (22%)  12/37 (32.4%)  5/37 (14%)  *p* = 0.26 | 8/22 (36%)  7/22 (32%)  4/22 (18%)  3/22 (14%) | 14/33 (42%)  13/33 (39%)  4/33 (12%)  2/33 (6%)  *p* = 0.68 | 10/25 (40%)  7/25 (28%)  6/25 (24%)  2/25 (8%) | 11/27 (41%)  12/27 (44%)  2/27 (7%)  2/27 (7%)  *p* = 0.36 |
| Avoid changing jobs  Yes  No  Don’t know | 22/110 (20.0%)  80/110 (72.7%)  8/110 (7.3%) | 10/30 (33%)  20/30 (67%)  2 | 16/38 (42%)  22/38 (58%)  *p* = 0.62 | 4/21 (19%)  17/21 (81%)  1 | 16/31 (52%)  15/31 (48%)  2  *p*= 0.02* | 7/24 (29%)  17/24 (71%)  1 | 9/26 (35%)  17/26 (65%)  1  *p* = 0.77 |
