## Supplemental Table 4 for "Multidimensional and Longitudinal Impact of a Genetic Diagnosis for Critically Ill Infants"

|  | **3 Months** | | **12 Months** | |
| --- | --- | --- | --- | --- |
| **Construct** | **Undiagnosed**  N = 38 | **Diagnosed**  N = 32 | **Undiagnosed**  N = 27 | **Diagnosed**  N = 25 |
| Importance | ß = 0.80 ± 0.37  ***p* = 0.04** | n/a | ß = 0.78 ± 0.60  *p =* 0.20 | n/a |
| Understand | ß = 0.40 ± 0.33  *p* = 0.23 | ß = 0.48 ± 0.43  *p* = 0.27 | ß = 0.35 ± 0.49  *p* = 0.48 | ß = 0.43 ± 0.46  *p* = 0.36 |
| Worry | ß = 0.73 ± 0.37  *p* = 0.06 | ß = 0.81 ± 0.42  *p =* 0.06 | ß = 0.14 ± 0.51  *p* = 0.79 | ß = 0.77 ± 0.49  *p =* 0.13 |
| Treatments | ß = 0.45 ± 0.29  *p* = 0.12 | ß = 0.40 ± 0.35  *p* = 0.27 | ß = 0.014 ± 0.51  *p* = 0.98 | ß = 0.98 ± 0.27  ***p* = 0.001** |
| Plan pregnancy | ß = -0.29 ± 0.41  *p* = 0.50 | ß = 0.08 ± 0.39  *p* = 0.83 | ß = -0.17 ± 0.48  *p =* 0.73 | ß = 0.77 ± 0.71  *p* = 0.30 |
| End of life | ß = 1.07 ± 0.41  ***p* = 0.013** | ß = 0.62 ± 1.10  *p* = 0.59 | ß = 0.61 ± 0.49  *p* = 0.22 | n/a |
| Reproductive | ß = -0.53 ± 0.36  *p* = 0.15 | ß = -0.25 ± 0.46  *p =* 0.58 | ß = -0.33 ± 0.41  *p* = 0.43 | ß = 0.13 ± 0.47  *p* = 0.78 |
| Prepare | ß = -0.12 ± 0.43  *p* = 0.78 | ß = 0.22 ± 0.47  *p =* 0.65 | ß = 0.01 ± 0.61  *p* = 0.98 | ß = 1.2 ± 0.35  ***p* = 0.002** |
| Alone | ß = 0.67 ± 0.34  *p* = 0.06 | ß = 0.58 ± 0.39  *p* = 0.15 | ß = 0.10 ± 0.48  *p* = 0.84 | ß = -0.19 ± 0.46  *p* = 0.69 |
| Connect | ß = -0.36 ± 0.34  *p* = 0.30 | ß = 0.14 ± 0.47  *p* = 0.77 | ß = -0.41 ± 0.52  *p* = 0.43 | ß = 0.74 ± 0.45  *p* = 0.11 |
| More guilt | ß = 0.61 ± 0.34  *p* = 0.08 | ß = 0.41 ± 0.45  *p* = 0.37 | ß = 0.61 ± 0.47  *p* = 0.21 | ß = -0.66 ± 0.38  *p* = 0.10 |

**Supplemental Table 4**

ß (± SEM) = change in Likert points for infants with metabolism consultation compared to genetics consultation.
