## Supplemental Table 3 for "Multidimensional and Longitudinal Impact of a Genetic Diagnosis for Critically Ill Infants"

| **Supplemental Table 3** | | | | |
| --- | --- | --- | --- | --- |
| **Undiagnosed** | | | | |
| **Domain** | **Baseline**  **(N = 110)** | **3 Months**  **(N = 38)** | **6 months**  **(N = 35)** | **12 months**  **(N = 27)** |
| *Importance*  Not at all  Slightly  Moderately  Very  Extremely | 1 (0.9%)  10 (9.1%)  18 (16.4%)  39 (35.5%)  42 (38.2%) | 3 (8%)  13 (34%)  10 (26%)  8 (21%)  4 (11%) | 4 (12.1%)  6 (18.2%)  10 (30.3%)  9 (27.2%)  4 (12.1%) | 5 (18.5%)  8 (29.6%)  4 (14.8%)  6 (22.2%)  4 (14.8%) |
| *Understanding*  Strongly disagree  Disagree  Neutral  Agree  Strongly agree | 0  1 (0.9%)  9 (8.2%)  31 (28.2%)  69 (62.7%) | 0  4 (10.5%)  4 (10.5%)  14 (36.8%)  16 (42.1%) | 1 (3.0%)  1 (3.0%)  5 (15.1%)  14 (42.4%)  12 (36.4%) | 1 (3.7%)  3 (11.1%)  5 (18.5%)  11 (40.7%)  7 (25.9%) |
| *Worry less*  Strongly disagree  Disagree  Neutral  Agree  Strongly agree | 2 (1.8%)  20 (18.2%)  28 (25.5%)  23 (20.9%)  37 (33.6%) | 1 (2.6%)  12 (31.6%)  10 (26.3%)  9 (23.7%)  6 (15.8%) | 1 (3.0%)  8 (24.2%)  10 (30.3%)  9 (27.2%)  5 (15.1%) | 1 (3.7%)  8 (29.5%)  7 (25.9%)  7 (25.9%)  4 (14.8%) |
| *Not help connect*  Strongly disagree  Disagree  Neutral  Agree  Strongly agree | 24 (21.8%)  44 (40.0%)  27 (24.5%)  10 (9.1%)  5 (4.5%) | 8 (21.1%)  15 (39.5%)  10 (26.3%)  4 (10.5%)  1 (2.6%) | 7 (21.2%)  14 (42.4%)  5 (15.1%)  4 (12.1%)  3 (9.1%) | 6 (22.2%)  11 (40.7%)  5 (18.5%)  3 (11.1%)  2 (7.4%) |
| *Feel more guilt*  Strongly disagree  Disagree  Neutral  Agree  Strongly agree | 29 (26.4%)  39 (35.5%)  22 (20.0%)  15 (13.6%)  5 (4.5%) | 7 (18.4%)  14 (36.8%)  9 (23.7%)  8 (21.1%)  0 | 8 (24.2%)  13 (39.3%)  5 (15.2%)  5 (15.2%)  2 (6.1%) | 5 (18.5%)  12 (44.4%)  5 (18.5%)  4 (14.8%)  1 (3.7%) |
| *Feel less alone*  Strongly disagree  Disagree  Neutral  Agree  Strongly agree | 10 (9.1%)  21 (19.1%)  38 (34.5%)  31 (28.2%)  9 (8.2%) | 2 (5.2%)  10 (26.3%)  12 (31.5%)  11 (28.9%)  3 (7.9%) | 1 (3.0%)  4 (12.1%)  15 (45.5%)  9 (27.3%)  4 (12.1%) | 2 (7.4%)  8 (29.6%)  9 (33.3%)  6 (22.2%)  2 (7.4%) |
| *Not help prepare*  Strongly disagree  Disagree  Neutral  Agree  Strongly agree | 38 (34.5%)  41 (37.3%)  10 (9.1%)  7 (6.4%)  14 (12.7%) | 13 (34.2%)  14 (36.8%)  4 (10.5%)  4 (10.5%)  3 (7.9%) | 6 (18.1%)  20 (60.6%)  3 (9.1%)  1 (3.0%)  3 (9.1%) | 10 (37.0%)  7 (25.9%)  5 (18.5%)  2 (7.4%)  3 (11.1%) |
| *Better treatments*  Strongly disagree  Disagree  Neutral  Agree  Strongly agree | 0  0  5 (4.5%)  49 (44.5%)  55 (50.0%) | 1 (2.6%)  1 (2.6%)  5 (13.2%)  21 (55.3%)  10 (26.3%) | 1 (3.0%)  1 (3.0%)  3 (9.1%)  16 (48.4%)  12 (36.4%) | 1 (3.7%)  3 (11.1%)  7 (25.9%)  8 (29.6%)  8 (29.6%) |
| *End-of-life plans*  Strongly disagree  Disagree  Neutral  Agree  Strongly agree | 6 (5.5%)  8 (7.3%)  27 (24.5%)  31 (28.2%)  34 (30.9%) | 4 (10.5%)  6 (15.8%)  8 (21.1%)  11 (28.9%)  9 (23.7%) | 4 (12.1%)  3 (9.1%)  9 (27.3%)  9 (27.3%)  8 (24.2%) | 2 (7.4%)  2 (7.4%)  11 (40.5%)  7 (25.9%)  5 (18.5%) |
| *Have more children*  Strongly disagree  Disagree  Neutral  Agree  Strongly agree | 11 (10.0%)  20 (18.2%)  24 (21.8%)  38 (34.5%)  17 (15.5%) | 1 (2.6%)  5 (13.2%)  6 (15.8%)  16 (42.1%)  10 (26.3%) | 2 (6.1%)  3 (9.1%)  4 (12.1%)  13 (39.4%)  11 (33.3%) | 0  2 (7.4%)  8 (29.6%)  10 (37.0%)  7 (25.9%) |
| *Pregnancy planning*  Strongly disagree  Disagree  Neutral  Agree  Strongly agree  N/A | 2 (1.8%)  3 (2.7%)  15 (13.6%)  36 (32.7%)  27 (24.5%)  27 (24.5%) | 0  1 (2.6%)  2 (5.2%)  9 (23.7%)  8 (21.1%)  18 (47.4%) | 1 (3.0%)  2 (6.0%)  0  9 (27.3%)  8 (24.2%)  13 (39.4%) | 0  0  4 (14.8%)  6 (22.2%)  6 (22.2%)  11 (40.7%) |

**Diagnosed**

| **Domain** | **3 Months**  **(N = 32)** | **6 months**  **(N = 23)** | **12 months (N=27)** |
| --- | --- | --- | --- |
| *Understanding*  Strongly disagree  Disagree  Neutral  Agree  Strongly agree | 1 (3.1%)  3 (9.4%)  5 (15.6%)  10 (31.3%)  13 (40.6%) | 1 (4.3%)  0  2 (8.7%)  9 (39.1%)  11 (47.8%) | 1 (3.7%)  6 (22.2%)  0  2 (7.4%)  16 (59.3%) |
| *Worry*  Strongly disagree  Disagree  Neutral  Agree  Strongly agree | 4 (12.5%)  8 (25.0%)  12 (37.5%)  5 (15.6%)  3 (9.4%) | 1 (4.3%)  8 (34.8%)  6 (26.1%)  7 (30.4%)  1 (4.3%) | 2 (7.4%)  11 (40.7%)  4 (14.8%)  5 (18.5%)  3 (11.1%) |
| *Feel less connection*  Strongly disagree  Disagree  Neutral  Agree  Strongly agree | 8 (25.0%)  6 (18.8%)  11 (34.4%)  5 (15.6%)  2 (6.3%) | 5 (21.7%)  11 (47.8%)  4 (17.4%)  2 (8.7%)  1 (4.3%) | 8 (29.6%)  4 (14.8%)  9 (33.0%)  4 (14.8%)  0 (0.0%) |
| *Feel more guilt*  Strongly disagree  Disagree  Neutral  Agree  Strongly agree | 5 (15.6%)  9 (28.1%)  8 (25.0%)  8 (25.0%)  2 (6.3%) | 7 (30.4%)  6 (26.1%)  6 (26.1%)  4 (17.4%)  0 | 6 (22.2%)  11 (40.7%)  5 (18.5%)  3 (11.1%)  0 (0.0%) |
| *Feel less alone*  Strongly disagree  Disagree  Neutral  Agree  Strongly agree | 1 (3.1%)  12 (37.5%)  7 (21.9%)  10 (31.3%)  2 (6.3%) | 3 (13.0%)  4 (17.4%)  6 (26.1%)  10 (43.4%)  0 | 2 (7.4%)  5 (18.5%)  8 (29.6%)  8 (29.6%)  2 (8.7%) |
| *Not help prepare*  Strongly disagree  Disagree  Neutral  Agree  Strongly agree | 9 (28.1%)  14 (43.8%)  4 (12.5%)  2 (6.3%)  3 (9.4%) | 9 (39.1%)  10 (43.5%)  2 (8.7%)  1 (4.3%)  1 (4.3%) | 9 (33.0%)  8 (29.6%)  6 (22.2%)  0  1 (3.7%) |
| *Treatments*  Strongly disagree  Disagree  Neutral  Agree  Strongly agree | 0  2 (6.3%)  5 (16.1%)  11 (35.5%)  13 (41.9%) | 0  0  5 (21.7%)  9 (39.1%)  9 (39.1%) | 0 (0.0%)  0 (0.0%)  7 (25.9%)  10 (37.0%)  8 (29.6%) |
| *End-of-life plans*  Strongly disagree  Disagree  Neutral  Agree  Strongly agree  N/A | 3 (9.5%)  0  4 (12.5%)  1 (3.1%)  2 (6.3%)  22 (68.8%) | 0  1 (4.3%)  4 (17.4%)  0  1 (4.3%)  17 (73.9%) | 1 (3.7%)  1 (3.7%)  4 (14.8%)  0 (0.0%)  0 (0.0%)  19 (70.4%) |
| *Have more children*  Strongly disagree  Disagree  Neutral  Agree  Strongly agree | 2 (6.3%)  6 (18.8%)  8 (25.0%)  10 (31.3%)  6 (18.8%) | 1 (4.3%)  4 (17.4%)  6 (26.1%)  6 (26.1%)  6 (26.1%) | 2 (7.4%)  3 (11.1%)  7 (25.9%)  10 (37.0%)  3 (11.1%) |
| *Family planning*  Strongly disagree  Disagree  Neutral  Agree  Strongly agree  N/A | 0  0  7 (21.9%)  7 (21.9%)  6 (18.8%)  12 (37.5%) | 0  4 (17.4%)  2 (8.7%)  6 (26.1%)  5 (21.7%)  6 (26.1%) | 1 (3.7%)  0 (0.0%)  5 (18.5%)  2 (7.4%)  7 (25.9%)  10 (37.0%) |
