## Supplemental Table 5 for "Multidimensional and Longitudinal Impact of a Genetic Diagnosis for Critically Ill Infants"

| **Supplemental Table 5** | | | | | | |
| --- | --- | --- | --- | --- | --- | --- |
|  | **3 Months** | | **6 Months** | | **12 Months** | |
| **Quality of Life Domain**  mean (SD) | **Undiagnosed**  N = 38 | **Diagnosed**  N = 32 | **Undiagnosed**  N = 33 | **Diagnosed**  N = 23 | **Undiagnosed**  N = 27 | **Diagnosed**  N = 25 |
| Overall Health | 60.9 (26.5) | 65.3 (27.0) | 71.8 (22.3) | 65.4 (24.8) | 81.9 (17.9) | 62.2 (28.8) |
|  |  | *p =* 0.50 |  | *p =* 0.33 |  | *p =* 0.006** |
| Physical activity | *not valid |  | *not valid |  | 83.5 (22.4) | 76.7 (26.7)  *p =* 0.33 |
| Growth and development | 78.0 (16.0) | 77.8 (22.0)  *p* = 0.96 | 80.3 (22.4) | 74.1 (22.6)  *p* = 0.32 | 81.7 (18.5) | 71.4 (25.1)  *p* = 0.10 |
| Discomfort and pain | 63.5 (23.0) | 74.2 (19.3)  *p* = 0.04** | 70.5 (20.2) | 68.5 (23.8)  *p* = 0.75 | 66.7 (22.7) | 73.0 (22.2)  *p* = 0.31 |
| Temperament and moods | 75.9 (15.2) | 75.7 (19.5)  *p* = 0.97 | 81.5 (10.9) | 83.0 (15.3)  *p* = 0.69 | 77.3 (14.0) | 80.8 (16.9)  *p* = 0.42 |
| Global behavior | *not valid |  | *not valid |  | 80.4 (20.8) | 88.4 (15.8)  *p* = 0.12 |
| Combined behavior | *not valid |  | *not valid |  | 63.0 (12.5) | 67.3 (9.5)  *p* = 0.17 |
| General health perception | 49.0 (21.8) | 49.1 (19.9)  *p* = 0.98 | 52.9 (15.8) | 47.3 (16.7)  *p* = 0.22 | 55.3 (16.7) | 43.0 (15.6)  *p* = 0.008** |
| Parental impact emotional | 60.0 (26.8) | 56.4 (29.1)  *p* = 0.60 | 71.8 (23.9) | 62.0 (27.6)  *p* = 0.17 | 69.7 (23.7) | 61.9 (26.7)  *p* = 0.28 |
| Parental impact time | 48.1 (20.7) | 52.7 (24.8)  *p* = 0.41 | 63.8 (15.4) | 53.4 (22.6)  *p* = 0.07 | 59.7 (20.4) | 56.0 (24.3)  *p* = 0.55 |
| Family cohesion | 76.6 (26.0) | 77.5 (24.1)  *p* = 0.88 | 79.1 (23.7) | 77.2 (27.6)  *p* = 0.79 | 77.0 (18.5) | 81.0 (21.2)  *p* = 0.48 |

Independent samples T test; mean (SD)

| **Supplemental Table 6** | | | | | | |
| --- | --- | --- | --- | --- | --- | --- |
|  | **3M***  **(N = 70)** | **6M***  **(N=56)** | **US Norms**  **2-11M**  **(N=247)** | **12M**  **(N=52)** | **US Norms****  **12-23M**  **(N=246)** | **US Norms ≥ 2 chronic conditions 4-71M****  **(N = 102)** |
| **Quality of Life Domain** |  |  |  |  |  |  |
| Physical activity | N/A | N/A | N/A | 80.2 (24.6) | 95.5 (17.13)  *p* < 0.001 | 85.2 (26.7)  *p* < 0.26 |
| Growth and development | 77.9 (18.8)  *p* < 0.001 | 77.8 (22.5)  *p* < 0.001 | 96.7 (8.0) | 76.7 (22.3) | 93.8 (14.3)  *p* < 0.001 | 86.0 (18.4)  *p =* 0.006 |
| Discomfort and pain | 68.4 (21.9)  *p =* 0.04 | 69.6 (21.6)  *p =* 0.12 | 73.6 (17.9) | 68.0 (23.5) | 76.9 (18.6)  *p =* 0.003 | 78.9 (19.1)  *p =* 0.006 |
| Temperament and moods | 75.8 (17.2)  *p* < 0.001 | 82.1 (12.8)  *p* < 0.001 | 83.4 (11.5) | 79.0 (15.4) | 82.7 (13.0)  *p =* 0.07 | 78.4 (16.3)  *p =* 0.002 |
| Combined behavior | N/A | N/A | N/A | 65.1 (11.3) | 79.4 (12.0)  *p* < 0.001 | 64.8 (18.0)  *p =* 0.91 |
| General health perception | 49.0 (20.8)  *p* < 0.001 | 50.6 (16.3)  *p* < 0.001 | 81.7 (13.0) | 49.4 (17.2) | 79.8 (13.5)  *p* < 0.001 | 65.5 (15.5)  *p* < 0.001 |
| Parental impact emotional | 58.4 (27.7)  *p* < 0.001 | 67.7 (25.7)  *p* < 0.001 | 92.7 (13.8) | 65.9 (25.3) | 89.9 (17.1)  *p* < 0.001 | 74.4 (24.8)  *p =* 0.048 |
| Parental impact time | 50.2 (22.6)  *p* < 0.001 | 59.7 (19.1)  *p* < 0.001 | 91.7 (17.7) | 57.9 (22.2) | 90.5 (20.3)  *p* < 0.001 | 82.0 (24.8)  *p* < 0.001 |
| Family cohesion | 77.0 (25.0)  *p =* 0.04 | 78.4 (25.1)  *p =* 0.14 | 82.6 (19.8) | 78.9 (19.8) | 78.9 (21.4)  *p* = 1.0 | 75.3 (24.0)  *p =* 0.35 |

***comparison to US Norms 2-11M

**comparison to 12M
