## Supplemental Figure 1 for "Multidimensional and Longitudinal Impact of a Genetic Diagnosis for Critically Ill Infants"

| **Figure S1. Follow-up survey items related to parent-perceived utility of a diagnosis.** | | |
| --- | --- | --- |
| *Undiagnosed infants* | *Diagnosed infants* | Response options |
| 1. How important is having a genetic diagnosis for your baby?  2. Having a genetic diagnosis will help me understand my baby's current health problems.  3. Having a genetic diagnosis will help me worry less about my baby's health problems.  4. Having a genetic diagnosis will not help me prepare for my baby's future.  5. Having a genetic diagnosis will help me find better treatments for my baby's health problems  6. Having a genetic diagnosis would help me make end-of-life decisions if my baby were at risk of dying.  7. Having a genetic diagnosis will not help me connect with other families  8. Having a genetic diagnosis will cause me to feel more guilt about my baby’s health problems  9. Having a genetic diagnosis will help me feel less alone  10. Having a genetic diagnosis will help me decide whether or not to have more children  11. Having a genetic diagnosis will help me plan for my/my partner’s next pregnancy [if applicable] | 1. [not asked]  2. Having a genetic diagnosis has helped me understand my baby's current health problems.  3. Having a genetic diagnosis has helped me worry less about my baby's health problems.  4. Having a genetic diagnosis has not helped me prepare for my baby's future.  5. Having a genetic diagnosis has helped me find better treatments for my baby's health problems  6. Having a genetic diagnosis has helped me to make end-of-life decisions for my baby [if applicable]  7. Having a genetic diagnosis has not helped me connect with other families  8. Having a genetic diagnosis has caused me to feel more guilt about my baby’s health problems  9. Having a genetic diagnosis has helped me feel less alone  10. Having a genetic diagnosis has helped me decide whether or not to have more children  11. Having a genetic diagnosis has helped me plan for my/my partner’s next pregnancy [if applicable] | 5-point Likert scale  [not at all important… extremely important]  5-point Likert scale  [strongly disagree… strongly agree] |
